# Assessing dose-response relations of lipid traits with coronary artery disease, all-cause mortality, and cause-specific mortality: a linear and non-linear Mendelian randomization study

**DOI:** 10.1101/2023.09.27.23296203

**Authors:** Guoyi Yang, Amy M Mason, Angela M Wood, C Mary Schooling, Stephen Burgess

## Abstract

**Importance:** Apolipoprotein B (apoB), low-density lipoprotein cholesterol (LDL-C), and possibly triglycerides (TG) play causal roles in the aetiology of coronary artery disease (CAD). However, trial evidence for effects of intensive LDL-C lowering and TG lowering on mortality is less definitive.

**Objectives:** To investigate dose-response relations of apoB, LDL-C, and TG with CAD and mortality overall, by sex, and by age.

**Design:** We performed linear Mendelian randomization (MR) analyses to assess the associations of genetically-predicted apoB, LDL-C, and TG with CAD, all-cause mortality, and cause-specific mortality. We also performed non-linear MR analyses, dividing the population into 10 strata, to assess stratum-specific estimates and characterise the shape of these associations.

**Setting:** UK Biobank.

**Participants:** 347,797 European ancestry participants (23,818 CAD cases and 23,848 deaths).

**Exposures:** Genetically-predicted apoB, LDL-C, and TG.

**Main outcomes and measures:** CAD, all-cause mortality, cardiovascular mortality, cancer mortality, and non-cardiovascular/cancer mortality.

**Results:** Genetically-predicted apoB was positively associated with CAD (odds ratio (OR) 1.65 per standard deviation increase [95% confidence interval 1.57, 1.73]), all-cause mortality (hazard ratio (HR) 1.11 [1.06, 1.16]), and cardiovascular mortality (HR 1.36 [1.24, 1.50]), with some evidence for stronger associations in men than women. Findings were similar for LDL-C. Genetically-predicted TG was positively associated with CAD (OR 1.60 [1.52, 1.69]), all-cause mortality (HR 1.08 [1.03, 1.13]), and cardiovascular mortality (HR 1.21 [1.09, 1.34]); however, sensitivity analyses suggested evidence of pleiotropy. The association of genetically-predicted TG with CAD persisted but its associations with mortality outcomes were attenuated towards the null after controlling for LDL-C.

Non-linear MR suggested the shapes of all these associations were monotonically increasing across the whole observed distribution of each lipid trait, with no diminution at low lipid levels. Such patterns were observed irrespective of sex or age.

**Conclusions and relevance:** Our findings suggest that apoB (or equivalently LDL-C) increases CAD risk, all-cause mortality, and cardiovascular mortality all in a dose-dependent way. TG likely increases CAD risk, although the possible presence of pleiotropy is a limitation. These insights highlight the importance of LDL-C lowering for reducing cardiovascular morbidity and mortality across its whole distribution.

**Key points:** 

**Question:** Do apolipoprotein B (apoB), low-density lipoprotein cholesterol (LDL-C), and triglycerides (TG) increase risk of coronary artery disease (CAD), all-cause mortality, or cause- specific mortality, and if so, what are the shapes of these relations?

**Findings:** In this Mendelian randomization study including 347,797 European ancestry participants from UK Biobank, genetically-predicted apoB and LDL-C were positively associated with CAD, all- cause mortality, and cardiovascular mortality all in a dose-dependent way. Genetically-predicted TG was positively associated with CAD, although the presence of pleiotropy was suggested.

**Meaning:** ApoB (or equivalently LDL-C) lowering reduces cardiovascular morbidity and mortality across its whole observed distribution.

## Introduction

Lipid management is essential for preventing cardiovascular morbidity and mortality.^1,2^ Apolipoprotein B (ApoB) is emerging as the predominant trait that accounts for the aetiological relations of lipid traits with coronary artery disease (CAD).^3^ A causal role of low-density lipoprotein cholesterol (LDL-C) in CAD is widely accepted,^4^ and a role of triglycerides (TG) is also gaining acceptance.^5,6^ However, dose-response relations of these lipid traits with CAD and mortality remain unclear.

Randomized controlled trials (RCTs) have shown LDL-C lowering reduces cardiovascular disease (CVD) events and mortality,^7^ and intensive LDL-C lowering further reduces CVD events compared with standard LDL-C lowering.^8–10^ However, no reduction in all-cause mortality or CVD mortality has been shown in trials comparing more intensive and less intensive statin therapy,^11–13^ or in trials adding PCSK9 inhibitors ^9^ or ezetimibe ^10^ to background statin therapy. Meta-regression of RCTs suggests the magnitude of LDL-C lowering is associated with greater reduction in all-cause mortality and CVD mortality in trials of people with higher baseline LDL-C.^14^ RCTs have shown therapies primarily lowering TG (i.e., fibrates and omega-3 supplements) reduce CVD events but have little detectable effect on all-cause mortality or CVD mortality.^15,16^ Taken together, these studies raise the question about the balance of risks and benefits for lipid lowering, particular for groups having low baseline LDL-C or TG, as well as for women, who have lower CVD risk than men, and for older people, who generally have more non-CVD comorbidities than younger people.

Observational studies have shown J-shaped relations of LDL-C and TG with all-cause mortality.^17,18^ However, the shape of these associations could also be an indicator of confounding or selection bias. Better understanding of the shape of causal relations of lipid traits with CAD and mortality has clinical implications for determining lipid lowering goals for CVD prevention.

Mendelian randomization (MR) takes advantage of genetic randomization at conception to obtain less confounded estimates than conventional observational studies.^19^ Previous MR studies have suggested apoB, LDL-C, and TG are positively associated with risk of CAD ^3,5,20^ and all-cause mortality.^21^ However, these studies did not take into account potential non-linearity and differences by sex or age.^3,5,20,21^ To address the gap, we firstly conducted linear MR analyses to assess the relations of genetically-predicted apoB, LDL-C, and TG with CAD, all-cause mortality, and cause-specific mortality. We then performed non-linear MR analyses to characterise the shape of these relations. We conducted subgroup analyses by sex because sex-specific effects are evident for LDL-C,^22^ TG,^23^ and some lipid modifiers.^24^ We also conducted subgroup analyses by age (<65 years and ≥65 years) because of potential concerns about efficacy of lipid lowering in older people.^25^

## Methods

### Study design

We performed linear and non-linear MR analyses in UK Biobank. UK Biobank recruited approximately 500,000 people (intended age 40-69 years, 94% self-reported European ancestry) from 2006 to 2010 in England, Scotland and Wales.^26^ Participants completed a range of physical assessments and questionnaires including socioeconomic characteristics, lifestyle, and health-related conditions, and provided samples for biological measurement and genotyping.^26^ Follow-up information was obtained by linking to national medical and mortality records (September 2021 update).^26^ We included 347,797 unrelated individuals of European ancestry with valid measurements of apoB, LDL-C and TG, and genomic data passing quality control as described previously.^27^

CAD was defined based on both prevalent cases (self-reported health conditions and medical records at recruitment) and incident events (International Classification of Diseases (ICD)-10 codes I20-I25) during the follow-up. All-cause mortality was determined by participant’s mortality status, and cause-specific mortality was divided into CVD mortality, cancer mortality, and non-CVD/cancer mortality using ICD-10 codes (eTable 1). We replicated the findings for all-cause mortality using parental mortality status from a meta-analysis of UK Biobank and LifeGen (*N*=1,012,240, 60% deceased) ^28^ to reduce selection bias and increase power.

### Genetic risk score for apoB, LDL-C, and TG

We extracted 163 independent (r^2^<0.001) genome-wide significant (*p* value <5×10^−8^) genetic instruments for apoB from a GWAS of the UK Biobank (http://www.nealelab.is/uk-biobank), and 313 for LDL-C and 373 for TG from European descent summary statistics in Global Lipids Genetics Consortium (GLGC) excluding the UK Biobank participants.^29^

We generated genetic risk scores (GRSs) for apoB, LDL-C, and TG respectively, weighted by variant-specific associations from the original GWAS. We calculated the proportion of variance explained by the GRS and the F-statistic to assess instrument strength. We also checked whether the GRSs were associated with possible confounders, i.e., Townsend deprivation index, current smoking, current alcohol drinking, and physical activity.

### Linear MR

We calculated linear MR estimates using the ratio method by dividing the association of the GRS with the outcome by the association of the GRS with the exposure. We transformed each lipid trait using inverse rank-normalization for comparability, and obtained genetic associations with apoB, LDL-C, and TG using linear regression. We obtained genetic associations with CAD using logistic regression, and with mortality outcomes using Cox proportional hazards regression. We used attained age (current age or age at death) as the time variable in Cox proportional hazards model. We adjusted for age, age^2^, sex, age * sex, age^2^ * sex, and the first 20 principal components for genetic associations. To assess the robustness of ratio estimates, we conducted sensitivity analyses using methods with different assumptions about instrumental validity, i.e., inverse-variance weighted (IVW),^30^ weighted median,^31^ MR Egger,^32^ and contamination mixture methods.^33^

We used multivariable MR to assess the association of each lipid trait controlling for potential pleiotropy.^34^ For each multivariable MR model, we combined all the genetic instruments, dropped duplicated SNPs, and removed correlated (r^2^≥0.001) SNPs. We used the remaining SNPs to generate GRSs for apoB, LDL-C, and TG respectively, taking weights from the original GWAS. Given the high correlation between apoB and LDL-C (r^2^=0.96), we adjusted for genetically-predicted TG in the calculation of apoB (and LDL-C) estimates, and adjusted for genetically-predicted LDL-C in the calculation of TG estimates. We calculated the conditional F-statistic to assess instrument strength to predict one trait conditional on the other trait in multivariable MR.^35^ We used multivariable IVW and multivariable MR Egger as sensitivity analyses. As further sensitivity analyses, we additionally included possible confounders associated with the GRS (*p* value <0.01) in multivariable MR analyses.

### Non-linear MR

We applied the fractional polynomial method to examine non-linear relations.^36^ We stratified the population into 10 strata using the doubly-ranked method, so that the instrument remains independent of confounders within strata.^37^ For each stratum of the population, a linear MR estimate was calculated using the ratio method as described above. We did not perform inverse rank-normalization transformation for each lipid trait in non-linear MR analyses, because RCTs suggest clinical benefits of statin therapy are determined by absolute reduction in LDL-C.^7^ We meta-regressed the linear MR estimates against the mean value of the exposure in each stratum.^36^ We used a trend test to assess whether a linear trend in the stratum-specific estimates exists, and a fractional polynomial test to examine whether a non-linear model fits the exposure-outcome relation better than a linear model.^36^

Differences by sex were assessed using a two-sided z-test.^38^ All statistical analyses were conducted using R version 4.2.1 and the packages “ieugwasr”, “MendelianRandomization”, and “SUMnlmr”.

### Ethical approval of studies and informed consent

This study has been conducted using the UK Biobank Resource (Application number 98032).

The UK Biobank obtained ethical approval from the North West Multi-centre Research Ethics Committee, and the participants provided written informed consent. The analysis of publicly available summary statistics does not require ethical approval.

## Results

### Baseline characteristics

Baseline characteristics of 347,797 participants included in this study are shown in Table 1. There were 23,818 people who developed CAD (including 17,136 prevalent cases), and 23,848 people who died. The GRSs explained 12.1%, 10.7%, and 14.8% of the variance in apoB, LDL-C, and TG, with F-statistics of 1835, 1610, and 2315 respectively. The GRSs for apoB and LDL-C were inversely associated with current smoking, and the GRS for TG was inversely associated with current alcohol drinking (eTable 2, *p* values 0.005, 0.002, and 0.001, respectively).

**Table 1.** Baseline characteristics of participants in the UK Biobank.

| Characteristics | UK Biobank (N=347,797) |
| --- | --- |
| Age at recruitment, years | 57.2 (8.0) |
| <65 years | 281,076 (80.8) |
| >=65 years | 66,721 (19.2) |
| Sex |  |
| Men | 159,467 (45.9) |
| Women | 188,330 (54.1) |
| ApoB, g/L | 1.0 (0.2) |
| LDL-C, mmol/L | 3.6 (0.9) |
| TG, mmol/L | 1.7 (1.0) |
| BMI, kg/m<sup>2</sup> | 27.4 (4.8) |
| SBP, mmHg | 137.6 (18.6) |
| Townsend deprivation index | -1.5 (3.0) |
| MET score, minutes/week | 2,662.6 (2710.6) |
| Smoking |  |
| Current | 35,727 (10.3) |
| Other | 312,070 (89.7) |
| Alcohol drinking |  |
| Current | 324,289 (93.2) |
| Other | 23,508 (6.8) |
| Lipid-lowering medication |  |
| Current | 58,851 (16.9) |
| Other | 288,946 (83.1) |
| CAD | 23,818 (6.8) |
| Deaths | 23,848 (6.9) |
| CVD deaths | 4,944 (1.4) |
| Cancer deaths | 12,247 (3.5) |
| Non-CVD/cancer deaths | 6,427 (1.8) |

a. ApoB, apolipoprotein B; BMI, body mass index; CAD, coronary artery disease; CVD, cardiovascular disease; LDL-C, low-density lipoprotein cholesterol; MET, metabolic equivalent task; SBP, systolic blood pressure; TG, triglycerides.
b. Data are mean (standard deviation) for continuous variables or *N* (%) for categorical variables.

### Linear MR analyses

In univariable MR, genetically-predicted apoB was positively associated with risk of CAD, all-cause mortality, and CVD mortality, with some evidence for stronger associations in men than women (Figure 1, *p* values for sex differences <0.001, 0.18, and 0.76, respectively). Genetically-predicted apoB had little association with cancer mortality, but had a positive association with non-CVD/cancer mortality in men and older people (Figure 1). Similar patterns were observed for LDL-C as for apoB (Figure 1). Genetically-predicted TG was positively associated with risk of CAD, all-cause mortality, CVD mortality, and non-CVD/cancer mortality (Figure 1). Sensitivity analyses using other analytic methods gave similar estimates for apoB and LDL-C; however, the MR Egger intercept indicated directional pleiotropy for TG on CAD, and estimates for TG were attenuated in weighted median and MR Egger methods (eTable 3).

**Figure 1.**
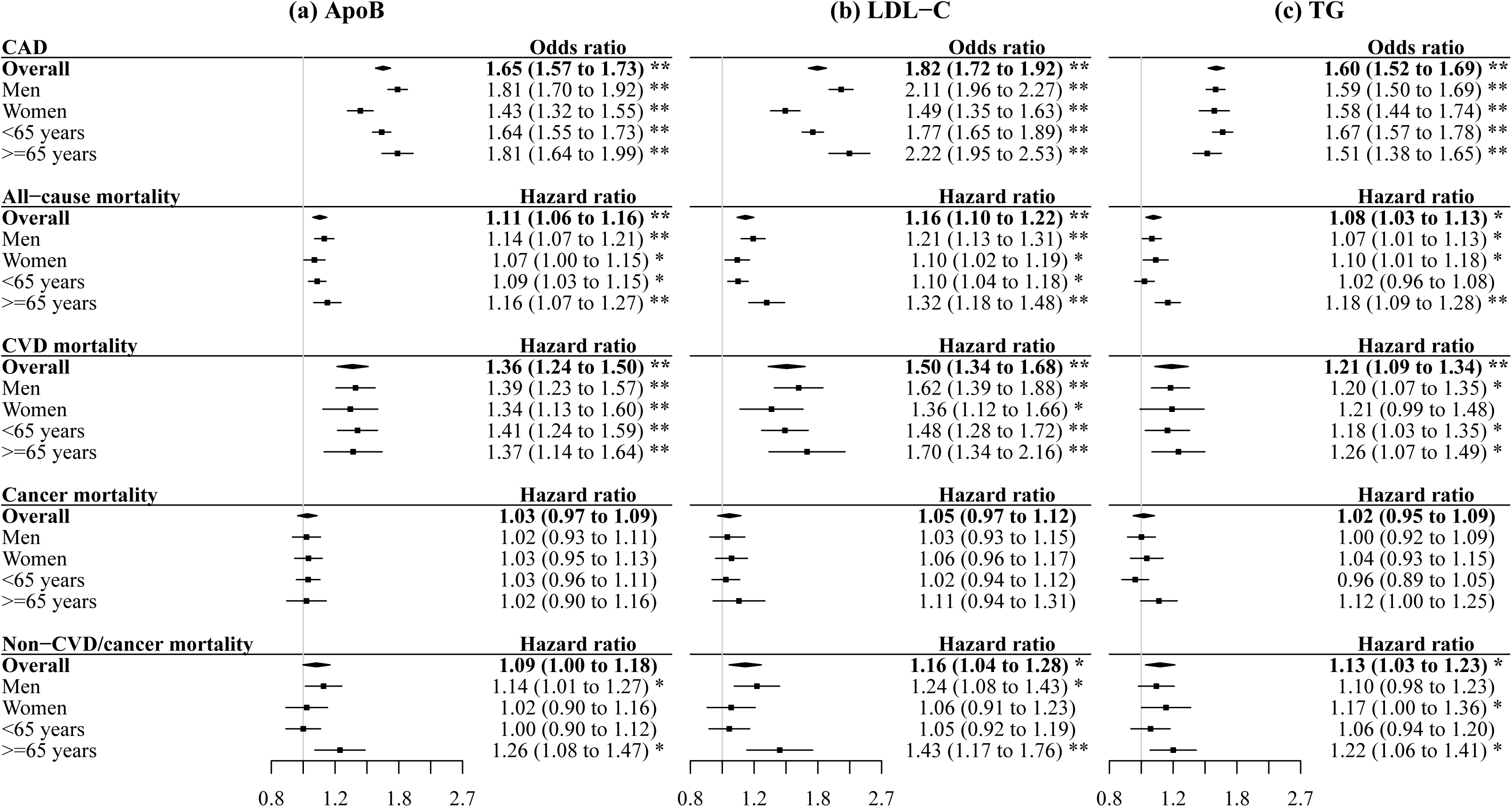
Univariable estimates for genetically-predicted lipid traits on CAD, all-cause mortality, and cause-specific mortality. a. ApoB, apolipoprotein B; CAD, coronary artery disease; CVD, cardiovascular disease; LDL-C, low-density lipoprotein cholesterol; TG, triglycerides. b. Estimates are expressed as odds ratio/hazard ratio (95% confidence interval) per standard deviation increase in genetically-predicted level of each lipid trait (approximately 0.24 g/L for apoB, 0.86 mmol/L for LDL-C, and 0.99 mmol/L for TG). c. * denotes *p* value <0.05; ** denotes *p* value <0.001.

In multivariable MR, the conditional F-statistics were 70, 51, and 61 for apoB, LDL-C, and TG, respectively. After controlling for TG, the associations of genetically-predicted apoB and LDL-C were similar to those in univariable MR (Figure 2). Although multivariable MR controlling for LDL-C showed some attenuation in the association of genetically-predicted TG with CAD, the positive association persisted (Figure 2). However, the associations of genetically-predicted TG with all-cause mortality, CVD mortality, and non-CVD/cancer mortality were attenuated towards the null after controlling for LDL-C (Figure 2). Similar results were obtained when using multivariable IVW and multivariable MR Egger, as well as additionally including possible confounders (i.e., current smoking for apoB and LDL-C, and current alcohol drinking for TG) (eTable 4).

**Figure 2.**
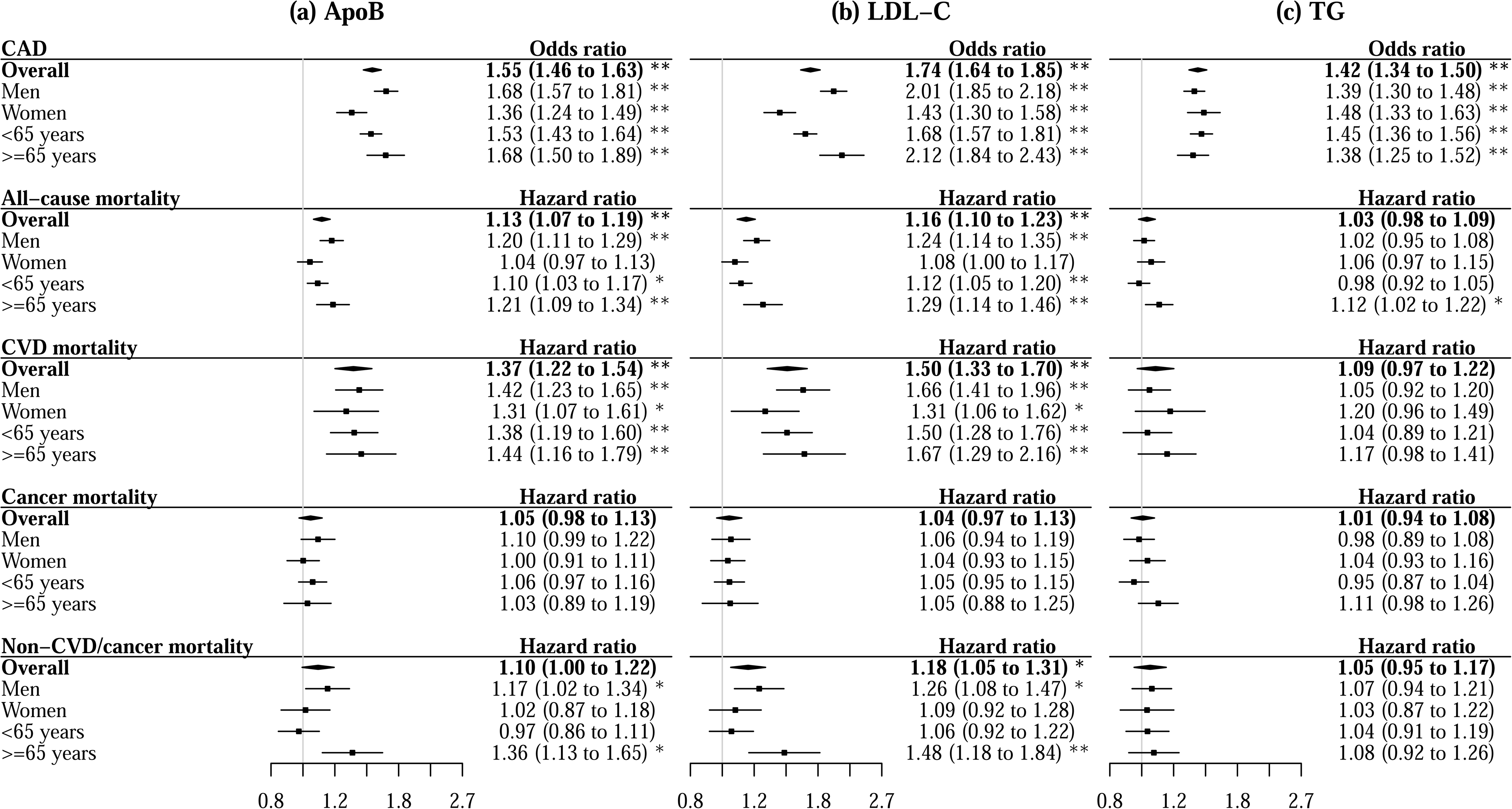
Multivariable estimates for genetically-predicted lipid traits on CAD, all-cause mortality, and cause-specific mortality. a. ApoB, apolipoprotein B; CAD, coronary artery disease; CVD, cardiovascular disease; LDL-C, low-density lipoprotein cholesterol; TG, triglycerides. b. Multivariable estimates for apoB and LDL-C were adjusted for TG, and multivariable estimates for TG were adjusted for LDL-C. c. Estimates are expressed as odds ratio/hazard ratio (95% confidence interval) per standard deviation increase in genetically-predicted level of each lipid trait (approximately 0.24 g/L for apoB, 0.86 mmol/L for LDL-C, and 0.99 mmol/L for TG). d. * denotes *p* value <0.05; ** denotes *p* value <0.001.

For replication using parental mortality status, genetically-predicted apoB, LDL-C, and TG were positively associated with parental all-cause mortality in univariable MR, although the MR Egger intercept indicated directional pleiotropy for TG (eTable 5). Multivariable MR controlling for TG gave similar results for apoB and LDL-C; however, multivariable MR controlling for LDL-C attenuated the association of TG with parental all-cause mortality (eTable 6).

### Non-linear MR analyses

We observed monotonically increasing associations of genetically-predicted apoB with CAD, all-cause mortality, and CVD mortality (Figure 3). Although non-linear MR suggested a weakening association of genetically-predicted apoB with CAD risk as apoB increased, stratum-specific associations remained positive across the whole distribution of apoB (eTable 7). There was no statistical evidence favoring a non-linear relation of genetically-predicted apoB with mortality outcomes over a linear one (Figure 3). Similar patterns were observed for LDL-C as for apoB (Figure 3, eTable 8). Genetically-predicted TG had monotonically increasing associations with CAD, all-cause mortality, CVD mortality, and non-CVD/cancer mortality (Figure 3). These associations were weakening with increasing TG, but stratum-specific associations were generally positive across the distribution of TG (eTable 9). Non-linear MR analyses by sex and age showed similar shapes of associations as in the whole population (eTable 10).

**Figure 3.**
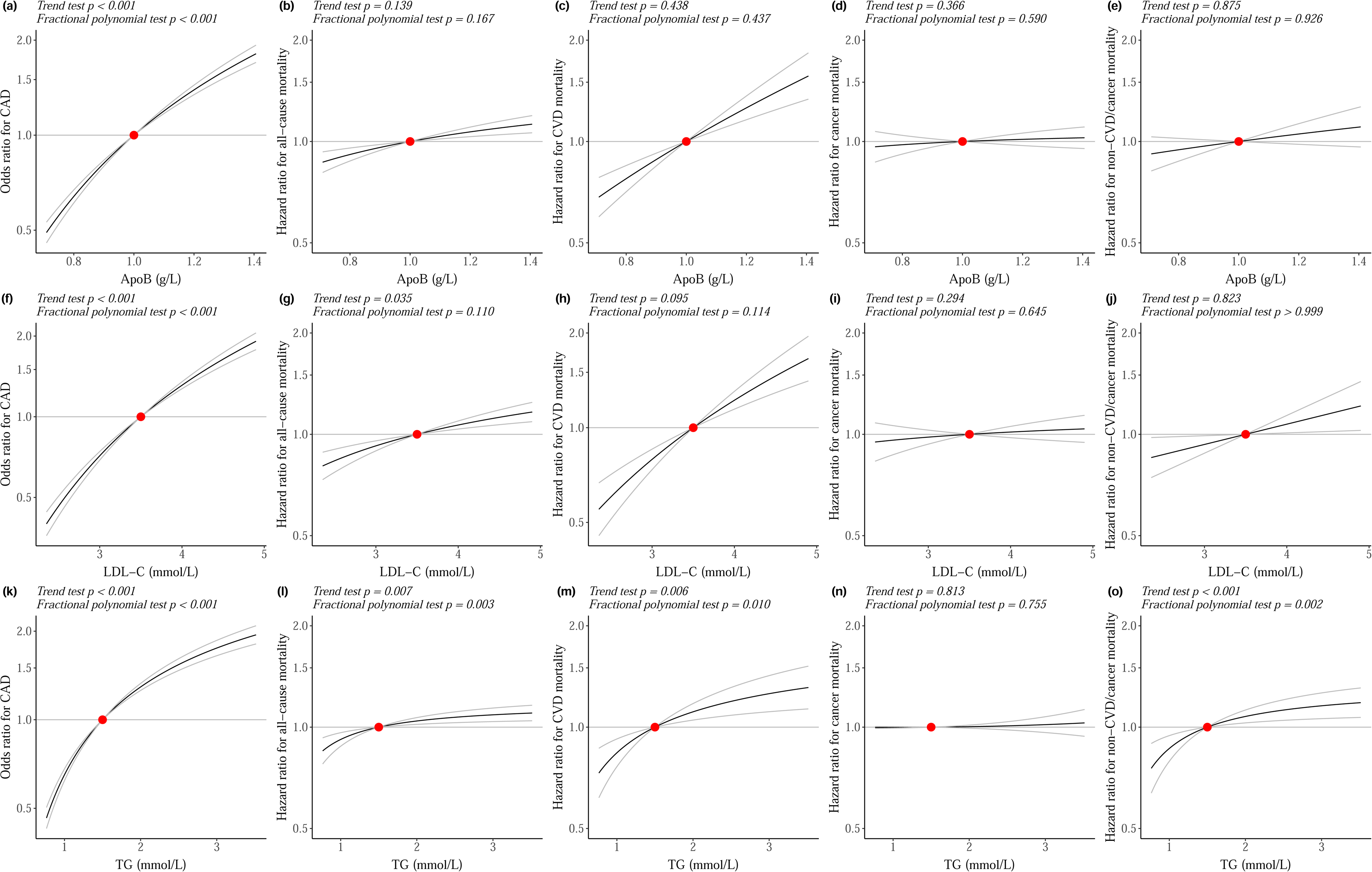
Shapes of the associations of genetically-predicted lipid traits with CAD, all-cause mortality, and cause-specific mortality. a. ApoB, apolipoprotein B; CAD, coronary artery disease; CVD, cardiovascular disease; LDL-C, low-density lipoprotein cholesterol; TG, triglycerides. b. The *x*-axis depicts apoB level in g/L, LDL-C level in mmol/L, or TG level in mmol/L. The *y*-axis depicts the odds ratio for CAD or hazard ratio for mortality outcomes with respect to the reference, plotted on a log scale. Reference is set to the median of each lipid trait (1.0 g/L for apoB, 3.5 mmol/L for LDL-C, and 1.5 mmol/L for TG). The black lines represent the dose-response relationship, and the grey lines represent the 95% confidence intervals. c. The trend test assesses whether a linear trend in the stratum-specific estimates exists, and the fractional polynomial test examines whether a non-linear model fits the exposure-outcome relationship better than a linear model.

## Discussion

Consistent with RCTs ^7,8,15,16^ and previous MR studies,^3,5,20,21^ we found positive associations of genetically-predicted apoB and LDL-C with CAD, all-cause mortality, and CVD mortality, and a possibly positive association of genetically-predicted TG with CAD. Our investigation has added to the evidence base by showing such associations hold across the whole distribution of apoB, LDL-C, or TG.

Our findings provide genetic evidence suggesting no threshold of lowering apoB or equivalently LDL-C (i.e., the main apoB-containing lipoprotein) for reducing risk of CAD, all-cause mortality, and CVD mortality, further supporting the concept of “the lower the better”. The retention of apoB-containing lipoproteins in the artery wall and subsequent release of cholesterol contents are essential for the initiation and progression of atherosclerosis.^39^ Our findings are consistent with RCTs showing intensive LDL-C lowering further reduces CVD events compared with standard LDL-C lowering.^8–10^ However, most RCTs failed to show significant incremental benefits of intensive LDL-C lowering on all-cause mortality or CVD mortality.^9–13^ This discrepancy may be related to insufficient difference in LDL-C between groups, short follow-up duration, and low proportion of death from CVD in RCTs,^9–13^ or differing effects on mortality between LDL-C lowering therapies.^40^ Notably, a trial has shown high-dose statin therapy further reduces all-cause mortality compared with low-dose therapy in Japanese with the mean LDL-C <90 mg/dL at baseline.^41^ Our findings are consistent with a recent MR study showing no threshold in the association of LDL-C with CAD, although their study used a different approach for instrument selection and lacked power to detect a significant association of LDL-C with all-cause mortality.^42^

The weakening association of genetically-predicted LDL-C with CAD risk as LDL-C increases should be interpreted cautiously, because fewer of those with higher LDL-C survived to be recruited ^43^ and more of them were depleted during follow-up,^44^ which may lead to an artefact of less harmful effects. Prevalent statin use could also mitigate the genetic effect of LDL-C on CAD risk, particularly in people with higher LDL-C.^45^ Furthermore, LDL-C confers greater absolute risk of CAD in people with higher LDL-C than people with lower LDL-C even for the same relative risk.

Despite consensus about the benefits of LDL-C lowering, previous MR studies showed LDL-C lowering may increase risk of type 2 diabetes ^46^ and some cancers.^47,48^ However, we found no adverse effects of LDL-C lowering on cancer mortality or non-cancer/CVD mortality, although we cannot exclude the possibility that LDL-C lowering may increase specific cancer risk. Consistently, a recent study showed long-term achievement of lower LDL-C levels, down to 20 mg/dL, is associated with lower CVD risk without significant safety concerns.^49^ Taken together, these findings suggest the cardiovascular benefits of LDL-C lowering outweigh its adverse effects, which results in a reduction in all-cause mortality, even in people with low baseline LDL-C.

Genetically-predicted LDL-C was more strongly associated with CAD and possibly all-cause mortality in men than women (*p* values for sex differences <0.001 and 0.07, respectively), which implies that men might benefit more from LDL-C lowering therapies than women. Our findings are consistent with a recent MR study showing sex-specific associations of LDL-C with CVD.^22^ However, meta-analysis of RCTs suggested similar beneficial effects of LDL-C lowering by statins on major CVD events and all-cause mortality for women and men,^7^ possibly attributed to under- representation of women (27%) in statin trials. Further investigations are warranted to understand potential factors underlying sex-specific effects of LDL-C, such as sex hormones.^50^

Our findings for TG are intriguing. In univariable MR, genetically-predicted TG was positively associated with risk of CAD, all-cause mortality, and CVD mortality. However, we found evidence of directional pleiotropy, which suggests potential bias or reflects different biological mechanisms involved in any effects of TG. In multivariable MR, which approximates the scenario for changing TG without altering LDL-C, the association of TG with CAD remained positive, but the associations with all-cause mortality and CVD mortality were attenuated towards the null. Correspondingly, RCTs show therapies primarily lowering TG (i.e., fibrates and omega-3 supplements) reduce CVD events but have little effect on mortality.^15,16^ It is also possible that TG lowering may affect CVD diagnosis, and thus its beneficial effect on CVD is an artefact mainly driven by reduction in soft events, such as revascularization.^51^

This is the first MR study comprehensively assessing relations of lipid traits with CAD, all-cause mortality, and cause-specific mortality, taking into account potential non-linearity as well as differences by sex and age. Nevertheless, this study has several limitations. First, MR relies on three rigorous assumptions of relevance, independence, and exclusion restriction, that is genetic instruments should be strongly related to the exposure, share no common cause with the outcome, and be independent of the outcome given the exposure.^19^ The GRSs were inversely associated with current smoking or alcohol drinking, which we addressed by using multivariable MR. Second, we used multivariable MR to assess the association of apoB (or equivalently LDL-C) adjusted for TG and vice versa, as they might have bi-directional relations. Adjustment for LDL-C might have inappropriately masked some effects of TG, because the clinical benefits of lowering TG could be driven by reduction in apoB ^52^ or by other mechanisms.^23^ Third, we conducted non-linear MR analyses to characterise the shapes of relations. However, due to a current lack of suitable methods, we were unable to perform non-linear multivariable MR analyses. Fourth, we obtained genetic instruments for apoB from the same study as genetic effects on CAD and mortality. Bias due to participant overlap depends on the strength of genetic associations with the exposure,^53^ and thus should not be substantial given the high F-statistic for apoB. Fifth, we calculated the GRSs for apoB, LDL-C, and TG from GWAS performed in a whole population and applied them to derive sex- and age-specific genetic associations. However, it is unlikely to have changed our results substantially given the genetics of most biomarkers are shared between women and men.^54^ Any minor discrepancies between overall estimates and the weighted average of subgroup estimates are likely due to non-collapsibility of odds ratio or hazard ratio. Sixth, the analyses were restricted to people of European ancestry and may not apply to other populations. Although we found monotonically increasing associations of LDL-C with CAD, all-cause mortality, and CVD mortality across the observed distribution of LDL-C, we cannot exclude the possibility of a threshold association in populations with relatively low LDL-C, such as East Asians.^55^ A previous MR study showed LDL-C is inversely associated with intracerebral haemorrhage in East Asians,^56^ which may detract from benefits of LDL-C lowering or could be due to selection bias. Finally, MR assesses lifelong effects of lipid traits, which cannot directly inform the quantitative effects of lipid-lowering therapies in the short term.

## Conclusions

This MR study suggests apoB (or equivalently LDL-C) increases risk of CAD, all-cause mortality, and CVD mortality all in a dose-dependent way. TG likely increases CAD risk, but the possible presence of pleiotropy is a limitation. These insights highlight the importance of LDL-C lowering for reducing CVD morbidity and mortality across its whole distribution.

## Supporting information

eTables 1-10

## Data Availability

This study has been conducted using the UK Biobank Resource under Application number 98032. Summary-level data analyzed are available in the website http://www.nealelab.is/uk-biobank/ for UK Biobank (Neale lab), http://csg.sph.umich.edu/willer/public/glgc-lipids2021/ for GLGC, and https://www.ebi.ac.uk/gwas/ for GWAS of parental all-cause mortality.

## Author contributions

GY and SB designed the study. GY undertook analyses with feedback from AMM, AMW, CMS and SB. GY drafted the manuscript with critical feedback and revisions from AMM, AMW, CMS and SB. All authors read and approved the final version of the manuscript.

## Conflicts of interest and financial disclosures

The authors declared no conflict of interest.

## Funding/support and role of funder/sponsor

GY is supported by the Bau Tsu Zung Bau Kwan Yeu Hing Research and Clinical Fellowship from the University of Hong Kong. AMM is funded by the EU/EFPIA Innovative Medicines Initiative Joint Undertaking BigData@Heart grant 116074. SB is supported by the Wellcome Trust (225790/Z/22/Z) and the United Kingdom Research and Innovation Medical Research Council (MC_UU_00002/7). This research was supported by British Heart Foundation (RG/13/13/30194; RG/18/13/33946), BHF Cambridge CRE (RE/18/1/34212), and the National Institute for Health Research Cambridge Biomedical Research Centre (NIHR203312). The views expressed are those of the authors and not necessarily those of the National Institute for Health Research or the Department of Health and Social Care. The funders had no role in the study design, analyses, or interpretation of results. For the purpose of open access, the author has applied a Creative Commons Attribution (CC BY) licence to any Author Accepted Manuscript version arising from this submission.

## Data access, responsibility, and analysis

GY had full access to all the data in the study and takes responsibility for the integrity of the data and the accuracy of the data analysis.

## Acknowledgements

The authors acknowledge the UK Biobank for approving our application, and the Neale lab, GLGC and Timmers PR, et al. for their publicly available summary data.

## Notes

### Competing Interest Statement

The authors have declared no competing interest.

### Author Declarations

This study has been conducted using the UK Biobank Resource (Application number 98032). The UK Biobank obtained ethical approval from the North West Multi-centre Research Ethics Committee, and the participants provided written informed consent. The analysis of publicly available summary statistics does not require ethical approval.

