## Supplementary material for "Assessing dose-response relations of lipid traits with coronary artery disease, all-cause mortality, and cause-specific mortality: a linear and non-linear Mendelian randomization study": eTables 1-10

**Online-Only Supplements**

**eTable 1.** International Classification of Diseases (ICD)-10 codes for cause-specific mortality outcomes.

**eTable 2.** Mendelian randomization estimates using the ratio method for genetically-predicted apoB, LDL-C, and TG on possible confounders.

**eTable 3.** Univariable Mendelian randomization estimates for genetically-predicted apoB, LDL-C, and TG on CAD, all-cause mortality, and cause-specific mortality.

**eTable 4.** Multivariable Mendelian randomization estimates for genetically-predicted apoB, LDL-C, and TG on CAD, all-cause mortality, and cause-specific mortality controlling for other lipid traits and possible confounders.

**eTable 5.** Univariable Mendelian randomization estimates for genetically-predicted apoB, LDL-C, and TG on parental all-cause mortality.

**eTable 6.** Multivariable Mendelian randomization estimates for genetically-predicted apoB, LDL-C, and TG on parental all-cause mortality controlling for other lipid traits and possible confounders.

**eTable 7.** Stratum-specific Mendelian randomization estimates for genetically-predicted apoB on CAD, all-cause mortality, and cause-specific mortality.

**eTable 8.** Stratum-specific Mendelian randomization estimates for genetically-predicted LDL-C on CAD, all-cause mortality, and cause-specific mortality.

**eTable 9.** Stratum-specific Mendelian randomization estimates for genetically-predicted TG on CAD, all-cause mortality, and cause-specific mortality.

**eTable 10.** Non-linear Mendelian randomization tests for genetically-predicted apoB, LDL-C, and TG on CAD, all-cause mortality, and cause-specific mortality overall, by sex, and by age.

**eTable 1. International Classification of Diseases (ICD)-10 codes for cause-specific mortality outcomes.**

| Cause-specific mortality | ICD-10 codes |
|---|---|
| CVD | G45, I01, I03-I82, I87, I95-I99, F01, Q20-Q28, R96 |
| Cancer | C00-C97, D00-D48 |
| Non-CVD/cancer | A00-A99, B00-B99, D50-D99,E00-E99, F00, F02-F99, G00-G44, G46-G99, H00-H99, I00, I02, I83-I86, I88-I89, J00-J99, K00-K99, L00-L99, M00-M99, N00-N99, O00-O99, P00-P99, Q00-Q18, Q30-Q99, S00-S99, T00-T99, U04, V00-V99, W00-W99, X00-X99, Y00-Y99, Z00-Z99 |

a. CVD, cardiovascular disease; ICD, International Classification of Diseases.

**eTable 2. Mendelian randomization estimates using the ratio method for genetically-predicted apoB, LDL-C, and TG on possible confounders.**

| Exposure | Outcome | Measure | Estimates | *P* value |
|---|---|---|---|---|
| ApoB | Townsend deprivation index | Beta | -0.04 (-0.07 to -0.005) | 0.025 |
| | MET score | Beta | 4.31 (-30.12 to 38.73) | 0.806 |
| | Current smoking | Odds ratio | 0.95 (0.91 to 0.98) | 0.005 |
| | Current alcohol drinking | Odds ratio | 0.98 (0.94 to 1.03) | 0.390 |
| LDL-C | Townsend deprivation index | Beta | -0.04 (-0.08 to 0.001) | 0.057 |
| | MET score | Beta | 25.77 (-15.28 to 66.83) | 0.219 |
| | Current smoking | Odds ratio | 0.93 (0.89 to 0.97) | 0.002 |
| | Current alcohol drinking | Odds ratio | 0.97 (0.92 to 1.02) | 0.247 |
| TG | Townsend deprivation index | Beta | 0.05 (0.01 to 0.08) | 0.013 |
| | MET score | Beta | 28.04 (-9.54 to 65.62) | 0.144 |
| | Current smoking | Odds ratio | 1.01 (0.97 to 1.05) | 0.617 |
| | Current alcohol drinking | Odds ratio | 0.92 (0.87 to 0.96) | 0.001 |

a. ApoB, apolipoprotein B; LDL-C, low-density lipoprotein cholesterol; MET, metabolic equivalent task; TG, triglycerides.

b. Estimates are expressed as beta/odds ratio (95% confidence interval) per standard deviation increase in each lipid trait.

**eTable 3. Univariable Mendelian randomization estimates for genetically-predicted apoB, LDL-C, and TG on CAD, all-cause mortality, and cause-specific mortality.**

| Exposure | Outcome | Method | Odds ratio/ Hazard ratio | *P* value | *P* value (Egger intercept) |
|---|---|---|---|---|---|
| ApoB | CAD | IVW | 1.66 (1.46 to 1.87) | <0.001 | |
| | CAD | Weighted median | 1.57 (1.42 to 1.74) | <0.001 | |
| | CAD | MR Egger | 1.74 (1.46 to 2.06) | <0.001 | 0.423 |
| | CAD | Conmix | 1.87 (1.68 to 2.01) | <0.001 | |
| | All-cause mortality | IVW | 1.11 (1.05 to 1.17) | <0.001 | |
| | All-cause mortality | Weighted median | 1.08 (1.00 to 1.17) | 0.059 | |
| | All-cause mortality | MR Egger | 1.12 (1.04 to 1.20) | 0.004 | 0.868 |
| | All-cause mortality | Conmix | 1.09 (1.03 to 1.14) | 0.007 | |
| | CVD mortality | IVW | 1.36 (1.20 to 1.55) | <0.001 | |
| | CVD mortality | Weighted median | 1.44 (1.22 to 1.69) | <0.001 | |
| | CVD mortality | MR Egger | 1.54 (1.28 to 1.84) | <0.001 | 0.065 |
| | CVD mortality | Conmix | 1.39 (1.25 to 1.58) | <0.001 | |
| | Cancer mortality | IVW | 1.03 (0.96 to 1.10) | 0.464 | |
| | Cancer mortality | Weighted median | 0.99 (0.90 to 1.09) | 0.807 | |
| | Cancer mortality | MR Egger | 0.96 (0.87 to 1.05) | 0.359 | 0.035 |
| | Cancer mortality | Conmix | 1.02 (0.93 to 1.09) | 0.744 | |
| | Non-CVD/cancer mortality | IVW | 1.10 (0.99 to 1.21) | 0.071 | |
| | Non-CVD/cancer mortality | Weighted median | 1.00 (0.87 to 1.16) | 0.956 | |
| | Non-CVD/cancer mortality | MR Egger | 1.19 (1.03 to 1.36) | 0.016 | 0.111 |
| | Non-CVD/cancer mortality | Conmix | 1.03 (0.90 to 1.12) | 0.757 | |
| LDL-C | CAD | IVW | 1.73 (1.55 to 1.93) | <0.001 | |
| | CAD | Weighted median | 1.53 (1.38 to 1.70) | <0.001 | |
| | CAD | MR Egger | 1.83 (1.57 to 2.12) | <0.001 | 0.271 |
| | CAD | Conmix | 1.71 (1.61 to 1.83) | <0.001 | |
| | All-cause mortality | IVW | 1.15 (1.08 to 1.21) | <0.001 | |
| | All-cause mortality | Weighted median | 1.12 (1.02 to 1.23) | 0.015 | |
| | All-cause mortality | MR Egger | 1.15 (1.06 to 1.24) | <0.001 | 0.961 |
| | All-cause mortality | Conmix | 1.12 (1.06 to 1.22) | 0.001 | |
| | CVD mortality | IVW | 1.47 (1.29 to 1.67) | <0.001 | |
| | CVD mortality | Weighted median | 1.63 (1.33 to 1.99) | <0.001 | |
| | CVD mortality | MR Egger | 1.56 (1.31 to 1.86) | <0.001 | 0.328 |
| | CVD mortality | Conmix | 1.45 (1.27 to 1.64) | <0.001 | |
| | Cancer mortality | IVW | 1.04 (0.96 to 1.12) | 0.296 | |
| | Cancer mortality | Weighted median | 0.99 (0.88 to 1.11) | 0.845 | |
| | Cancer mortality | MR Egger | 1.00 (0.90 to 1.10) | 0.952 | 0.206 |
| | Cancer mortality | Conmix | 1.02 (0.96 to 1.09) | 0.528 | |
| | Non-CVD/cancer mortality | IVW | 1.15 (1.03 to 1.29) | 0.014 | |
| | Non-CVD/cancer mortality | Weighted median | 1.08 (0.92 to 1.28) | 0.336 | |
| | Non-CVD/cancer mortality | MR Egger | 1.19 (1.03 to 1.39) | 0.020 | 0.461 |

| | | | | | |
|---|---|---|---|---|---|
| | Non-CVD/cancer mortality | Conmix | 1.08 (0.98 to 1.21) | 0.106 | |
| TG | CAD | IVW | 1.59 (1.47 to 1.73) | <0.001 | |
| | CAD | Weighted median | 1.49 (1.34 to 1.65) | <0.001 | |
| | CAD | MR Egger | 1.37 (1.23 to 1.53) | <0.001 | <0.001 |
| | CAD | Conmix | 1.65 (1.51 to 1.81) | <0.001 | |
| | All-cause mortality | IVW | 1.07 (1.01 to 1.13) | 0.016 | |
| | All-cause mortality | Weighted median | 1.00 (0.92 to 1.08) | 0.942 | |
| | All-cause mortality | MR Egger | 1.03 (0.95 to 1.11) | 0.522 | 0.142 |
| | All-cause mortality | Conmix | 1.04 (1.00 to 1.10) | 0.033 | |
| | CVD mortality | IVW | 1.19 (1.06 to 1.34) | 0.003 | |
| | CVD mortality | Weighted median | 1.14 (0.95 to 1.36) | 0.152 | |
| | CVD mortality | MR Egger | 1.08 (0.91 to 1.27) | 0.377 | 0.105 |
| | CVD mortality | Conmix | 1.19 (1.08 to 1.33) | <0.001 | |
| | Cancer mortality | IVW | 1.02 (0.95 to 1.10) | 0.570 | |
| | Cancer mortality | Weighted median | 0.98 (0.88 to 1.09) | 0.707 | |
| | Cancer mortality | MR Egger | 0.98 (0.89 to 1.09) | 0.765 | 0.330 |
| | Cancer mortality | Conmix | 1.01 (0.95 to 1.07) | 0.729 | |
| | Non-CVD/cancer mortality | IVW | 1.09 (0.99 to 1.22) | 0.090 | |
| | Non-CVD/cancer mortality | Weighted median | 0.94 (0.81 to 1.10) | 0.468 | |
| | Non-CVD/cancer mortality | MR Egger | 1.08 (0.93 to 1.25) | 0.323 | 0.784 |
| | Non-CVD/cancer mortality | Conmix | 1.02 (0.93 to 1.12) | 0.607 | |

a. ApoB, apolipoprotein B; CAD, coronary artery disease; Conmix, contamination mixture method; CVD, cardiovascular disease; IVW, inverse-variance weighted; LDL-C, low-density lipoprotein cholesterol; TG, triglycerides.

b. Estimates are expressed as odds ratio (95% confidence interval) for CAD, or hazard ratio (95% confidence interval) for mortality outcomes per standard deviation increase in each lipid trait.

**eTable 4. Multivariable Mendelian randomization estimates for genetically-predicted apoB, LDL-C, and TG on CAD, all-cause mortality, and cause-specific mortality controlling for other lipid traits and possible confounders.**

| Exposure | Adjusted for | Outcome | Method | Odds ratio/Hazard ratio | *P* value | *P* value (Egger intercept) |
|---|---|---|---|---|---|---|
| ApoB | TG | CAD | IVW | 1.58 (1.43 to 1.75) | <0.001 | |
| ApoB | TG | CAD | MR Egger | 1.55 (1.39 to 1.74) | <0.001 | 0.478 |
| ApoB | TG and smoking | CAD | IVW | 1.60 (1.45 to 1.76) | <0.001 | |
| ApoB | TG and smoking | CAD | MR Egger | 1.57 (1.41 to 1.76) | <0.001 | 0.565 |
| ApoB | TG | All-cause mortality | IVW | 1.13 (1.06 to 1.20) | <0.001 | |
| ApoB | TG | All-cause mortality | MR Egger | 1.11 (1.04 to 1.19) | 0.002 | 0.208 |
| ApoB | TG and smoking | All-cause mortality | IVW | 1.14 (1.08 to 1.21) | <0.001 | |
| ApoB | TG and smoking | All-cause mortality | MR Egger | 1.13 (1.05 to 1.20) | <0.001 | 0.301 |
| ApoB | TG | CVD mortality | IVW | 1.40 (1.23 to 1.59) | <0.001 | |
| ApoB | TG | CVD mortality | MR Egger | 1.39 (1.21 to 1.61) | <0.001 | 0.879 |
| ApoB | TG and smoking | CVD mortality | IVW | 1.40 (1.23 to 1.59) | <0.001 | |
| ApoB | TG and smoking | CVD mortality | MR Egger | 1.39 (1.21 to 1.61) | <0.001 | 0.889 |
| ApoB | TG | Cancer mortality | IVW | 1.05 (0.97 to 1.13) | 0.263 | |
| ApoB | TG | Cancer mortality | MR Egger | 1.02 (0.93 to 1.11) | 0.733 | 0.123 |
| ApoB | TG and smoking | Cancer mortality | IVW | 1.06 (0.98 to 1.14) | 0.161 | |
| ApoB | TG and smoking | Cancer mortality | MR Egger | 1.03 (0.94 to 1.12) | 0.507 | 0.170 |
| ApoB | TG | Non-CVD/cancer mortality | IVW | 1.12 (1.00 to 1.25) | 0.046 | |
| ApoB | TG | Non-CVD/cancer mortality | MR Egger | 1.11 (0.98 to 1.26) | 0.089 | 0.822 |
| ApoB | TG and smoking | Non-CVD/cancer mortality | IVW | 1.14 (1.02 to 1.27) | 0.020 | |
| ApoB | TG and smoking | Non-CVD/cancer mortality | MR Egger | 1.14 (1.01 to 1.29) | 0.037 | 0.989 |
| LDL-C | TG | CAD | IVW | 1.64 (1.46 to 1.84) | <0.001 | |
| LDL-C | TG | CAD | MR Egger | 1.75 (1.53 to 2.00) | <0.001 | 0.082 |
| LDL-C | TG and smoking | CAD | IVW | 1.65 (1.47 to 1.85) | <0.001 | |
| LDL-C | TG and smoking | CAD | MR Egger | 1.77 (1.54 to 2.03) | <0.001 | 0.068 |

| | | | | | | |
|---|---|---|---|---|---|---|
| LDL-C | TG | All-cause mortality | IVW | 1.15 (1.08 to 1.22) | <0.001 | |
| LDL-C | TG | All-cause mortality | MR Egger | 1.14 (1.06 to 1.23) | 0.001 | 0.765 |
| LDL-C | TG and smoking | All-cause mortality | IVW | 1.16 (1.09 to 1.24) | <0.001 | |
| LDL-C | TG and smoking | All-cause mortality | MR Egger | 1.16 (1.07 to 1.25) | <0.001 | 0.893 |
| LDL-C | TG | CVD mortality | IVW | 1.44 (1.26 to 1.66) | <0.001 | |
| LDL-C | TG | CVD mortality | MR Egger | 1.51 (1.29 to 1.78) | <0.001 | 0.301 |
| LDL-C | TG and smoking | CVD mortality | IVW | 1.45 (1.26 to 1.66) | <0.001 | |
| LDL-C | TG and smoking | CVD mortality | MR Egger | 1.52 (1.29 to 1.79) | <0.001 | 0.294 |
| LDL-C | TG | Cancer mortality | IVW | 1.04 (0.96 to 1.13) | 0.311 | |
| LDL-C | TG | Cancer mortality | MR Egger | 1.01 (0.91 to 1.11) | 0.876 | 0.197 |
| LDL-C | TG and smoking | Cancer mortality | IVW | 1.05 (0.97 to 1.14) | 0.220 | |
| LDL-C | TG and smoking | Cancer mortality | MR Egger | 1.02 (0.92 to 1.12) | 0.695 | 0.238 |
| LDL-C | TG | Non-CVD/cancer mortality | IVW | 1.17 (1.04 to 1.33) | 0.010 | |
| LDL-C | TG | Non-CVD/cancer mortality | MR Egger | 1.18 (1.02 to 1.37) | 0.024 | 0.874 |
| LDL-C | TG and smoking | Non-CVD/cancer mortality | IVW | 1.19 (1.06 to 1.35) | 0.005 | |
| LDL-C | TG and smoking | Non-CVD/cancer mortality | MR Egger | 1.21 (1.04 to 1.40) | 0.011 | 0.769 |
| TG | LDL-C | CAD | IVW | 1.31 (1.18 to 1.46) | <0.001 | |
| TG | LDL-C | CAD | MR Egger | 1.25 (1.09 to 1.43) | 0.001 | 0.277 |
| TG | LDL-C and alcohol drinking | CAD | IVW | 1.30 (1.18 to 1.45) | <0.001 | |
| TG | LDL-C and alcohol drinking | CAD | MR Egger | 1.26 (1.10 to 1.44) | 0.001 | 0.442 |
| TG | LDL-C | All-cause mortality | IVW | 1.01 (0.95 to 1.07) | 0.714 | |
| TG | LDL-C | All-cause mortality | MR Egger | 0.96 (0.89 to 1.04) | 0.335 | 0.048 |
| TG | LDL-C and alcohol drinking | All-cause mortality | IVW | 1.01 (0.95 to 1.07) | 0.794 | |
| TG | LDL-C and alcohol drinking | All-cause mortality | MR Egger | 0.97 (0.90 to 1.04) | 0.393 | 0.094 |
| TG | LDL-C | CVD mortality | IVW | 1.04 (0.92 to 1.18) | 0.567 | |
| TG | LDL-C | CVD mortality | MR Egger | 0.97 (0.83 to 1.14) | 0.750 | 0.227 |
| TG | LDL-C and alcohol drinking | CVD mortality | IVW | 1.03 (0.91 to 1.17) | 0.600 | |
| TG | LDL-C and alcohol drinking | CVD mortality | MR Egger | 0.98 (0.83 to 1.15) | 0.789 | 0.285 |
| TG | LDL-C | Cancer mortality | IVW | 1.01 (0.93 to 1.09) | 0.828 | |

| | | | | | | |
|---|---|---|---|---|---|---|
| TG | LDL-C | Cancer mortality | MR Egger | 0.95 (0.86 to 1.05) | 0.313 | 0.061 |
| TG | LDL-C and alcohol drinking | Cancer mortality | IVW | 1.01 (0.93 to 1.08) | 0.885 | |
| TG | LDL-C and alcohol drinking | Cancer mortality | MR Egger | 0.95 (0.87 to 1.05) | 0.350 | 0.097 |
| TG | LDL-C | Non-CVD/cancer mortality | IVW | 1.00 (0.90 to 1.12) | 0.946 | |
| TG | LDL-C | Non-CVD/cancer mortality | MR Egger | 0.99 (0.85 to 1.14) | 0.858 | 0.713 |
| TG | LDL-C and alcohol drinking | Non-CVD/cancer mortality | IVW | 1.00 (0.89 to 1.12) | 0.997 | |
| TG | LDL-C and alcohol drinking | Non-CVD/cancer mortality | MR Egger | 0.99 (0.86 to 1.15) | 0.927 | 0.888 |

a. ApoB, apolipoprotein B; CAD, coronary artery disease; CVD, cardiovascular disease; IVW, inverse-variance weighted; LDL-C, low-density lipoprotein cholesterol; TG, triglycerides.

b. Estimates are expressed as odds ratio (95% confidence interval) for CAD, or hazard ratio (95% confidence interval) for mortality outcomes per standard deviation increase in each lipid trait.

**eTable 5. Univariable Mendelian randomization estimates for genetically-predicted apoB, LDL-C, and TG on parental all-cause mortality.**

| **Exposure** | **Method** | **Hazard ratio** | ***P* value** | ***P* value (Egger intercept)** |
|---|---|---|---|---|
| ApoB | IVW | 1.16 (1.12 to 1.21) | <0.001 | |
| | Weighted median | 1.15 (1.10 to 1.19) | <0.001 | |
| | MR Egger | 1.21 (1.14 to 1.27) | <0.001 | 0.081 |
| | Conmix | 1.17 (1.14 to 1.19) | <0.001 | |
| LDL-C | IVW | 1.18 (1.13 to 1.23) | <0.001 | |
| | Weighted median | 1.15 (1.10 to 1.20) | <0.001 | |
| | MR Egger | 1.19 (1.13 to 1.26) | <0.001 | 0.494 |
| | Conmix | 1.16 (1.12 to 1.19) | <0.001 | |
| TG | IVW | 1.14 (1.10 to 1.17) | <0.001 | |
| | Weighted median | 1.11 (1.07 to 1.15) | <0.001 | |
| | MR Egger | 1.08 (1.03 to 1.13) | 0.001 | 0.003 |
| | Conmix | 1.14 (1.12 to 1.16) | <0.001 | |

a. ApoB, apolipoprotein B; Conmix, contamination mixture method; IVW, inverse-variance weighted; LDL-C, low-density lipoprotein cholesterol; TG, triglycerides.

b. Estimates are expressed as hazard ratio (95% confidence interval) per standard deviation increase in each lipid trait.

**eTable 6. Multivariable Mendelian randomization estimates for genetically-predicted apoB, LDL-C, and TG on parental all-cause mortality controlling for other lipid traits and possible confounders.**

| Exposure | Adjusted for | Method | Hazard ratio | *P* value | *P* value (Egger intercept) |
|---|---|---|---|---|---|
| ApoB | TG | IVW | 1.17 (1.12 to 1.21) | <0.001 | |
| ApoB | TG | MR Egger | 1.17 (1.12 to 1.23) | <0.001 | 0.574 |
| ApoB | TG and smoking | IVW | 1.17 (1.13 to 1.22) | <0.001 | |
| ApoB | TG and smoking | MR Egger | 1.18 (1.13 to 1.23) | <0.001 | 0.434 |
| LDL-C | TG | IVW | 1.16 (1.11 to 1.21) | <0.001 | |
| LDL-C | TG | MR Egger | 1.19 (1.13 to 1.25) | <0.001 | 0.056 |
| LDL-C | TG and smoking | IVW | 1.16 (1.11 to 1.21) | <0.001 | |
| LDL-C | TG and smoking | MR Egger | 1.19 (1.14 to 1.25) | <0.001 | 0.042 |
| TG | LDL-C | IVW | 1.06 (1.02 to 1.10) | 0.003 | |
| TG | LDL-C | MR Egger | 1.03 (0.99 to 1.08) | 0.166 | 0.134 |
| TG | LDL-C and alcohol drinking | IVW | 1.06 (1.02 to 1.09) | 0.004 | |
| TG | LDL-C and alcohol drinking | MR Egger | 1.04 (0.99 to 1.09) | 0.134 | 0.235 |

a. ApoB, apolipoprotein B; IVW, inverse-variance weighted; LDL-C, low-density lipoprotein cholesterol; TG, triglycerides.

b. Estimates are expressed as hazard ratio (95% confidence interval) per standard deviation increase in each lipid trait.

**eTable 7. Stratum-specific Mendelian randomization estimates for genetically-predicted apoB on CAD, all-cause mortality, and cause-specific mortality.**

| Stratum | Mean apoB (g/L) | Outcome | Odds ratio/Hazard ratio | *P* value |
|---|---|---|---|---|
| 1 | 0.71 | CAD | 1.96 (1.75 to 2.21) | <0.001 |
| 2 | 0.81 | CAD | 2.28 (2.00 to 2.61) | <0.001 |
| 3 | 0.88 | CAD | 1.96 (1.70 to 2.27) | <0.001 |
| 4 | 0.94 | CAD | 1.70 (1.45 to 1.99) | <0.001 |
| 5 | 1.00 | CAD | 1.44 (1.21 to 1.71) | <0.001 |
| 6 | 1.05 | CAD | 1.36 (1.13 to 1.63) | 0.001 |
| 7 | 1.11 | CAD | 1.33 (1.10 to 1.60) | 0.003 |
| 8 | 1.18 | CAD | 1.44 (1.19 to 1.74) | <0.001 |
| 9 | 1.26 | CAD | 1.46 (1.21 to 1.77) | <0.001 |
| 10 | 1.41 | CAD | 1.20 (1.00 to 1.43) | 0.048 |
| 1 | 0.71 | All-cause mortality | 1.19 (1.05 to 1.34) | 0.008 |
| 2 | 0.81 | All-cause mortality | 1.11 (0.97 to 1.27) | 0.144 |
| 3 | 0.88 | All-cause mortality | 1.16 (1.01 to 1.34) | 0.039 |
| 4 | 0.94 | All-cause mortality | 1.12 (0.96 to 1.29) | 0.148 |
| 5 | 1.00 | All-cause mortality | 0.97 (0.83 to 1.12) | 0.642 |
| 6 | 1.05 | All-cause mortality | 1.06 (0.91 to 1.23) | 0.429 |
| 7 | 1.11 | All-cause mortality | 1.12 (0.96 to 1.30) | 0.164 |
| 8 | 1.18 | All-cause mortality | 1.28 (1.10 to 1.49) | 0.002 |
| 9 | 1.26 | All-cause mortality | 1.01 (0.87 to 1.18) | 0.851 |
| 10 | 1.41 | All-cause mortality | 1.01 (0.87 to 1.17) | 0.937 |
| 1 | 0.71 | CVD mortality | 1.29 (1.00 to 1.67) | 0.046 |
| 2 | 0.81 | CVD mortality | 1.64 (1.23 to 2.19) | 0.001 |
| 3 | 0.88 | CVD mortality | 1.47 (1.08 to 1.99) | 0.014 |
| 4 | 0.94 | CVD mortality | 1.71 (1.21 to 2.41) | 0.002 |
| 5 | 1.00 | CVD mortality | 1.08 (0.77 to 1.52) | 0.655 |
| 6 | 1.05 | CVD mortality | 1.35 (0.96 to 1.89) | 0.084 |
| 7 | 1.11 | CVD mortality | 1.07 (0.75 to 1.54) | 0.705 |
| 8 | 1.18 | CVD mortality | 1.45 (1.01 to 2.08) | 0.046 |
| 9 | 1.26 | CVD mortality | 1.18 (0.84 to 1.66) | 0.334 |
| 10 | 1.41 | CVD mortality | 1.34 (0.96 to 1.88) | 0.086 |
| 1 | 0.71 | Cancer mortality | 1.09 (0.90 to 1.32) | 0.369 |
| 2 | 0.81 | Cancer mortality | 0.97 (0.80 to 1.18) | 0.755 |
| 3 | 0.88 | Cancer mortality | 1.15 (0.94 to 1.41) | 0.174 |
| 4 | 0.94 | Cancer mortality | 0.96 (0.78 to 1.18) | 0.693 |
| 5 | 1.00 | Cancer mortality | 0.98 (0.79 to 1.20) | 0.823 |
| 6 | 1.05 | Cancer mortality | 0.98 (0.80 to 1.21) | 0.874 |
| 7 | 1.11 | Cancer mortality | 1.07 (0.87 to 1.32) | 0.520 |
| 8 | 1.18 | Cancer mortality | 1.19 (0.97 to 1.47) | 0.094 |
| 9 | 1.26 | Cancer mortality | 0.92 (0.75 to 1.12) | 0.388 |
| 10 | 1.41 | Cancer mortality | 0.96 (0.78 to 1.16) | 0.653 |

| | | | | |
|---|---|---|---|---|
| 1 | 0.71 | Non-CVD/cancer mortality | 1.20 (0.96 to 1.49) | 0.117 |
| 2 | 0.81 | Non-CVD/cancer mortality | 0.99 (0.77 to 1.28) | 0.965 |
| 3 | 0.88 | Non-CVD/cancer mortality | 0.97 (0.74 to 1.27) | 0.806 |
| 4 | 0.94 | Non-CVD/cancer mortality | 1.07 (0.81 to 1.42) | 0.639 |
| 5 | 1.00 | Non-CVD/cancer mortality | 0.83 (0.63 to 1.10) | 0.204 |
| 6 | 1.05 | Non-CVD/cancer mortality | 1.01 (0.75 to 1.35) | 0.965 |
| 7 | 1.11 | Non-CVD/cancer mortality | 1.26 (0.93 to 1.71) | 0.142 |
| 8 | 1.18 | Non-CVD/cancer mortality | 1.38 (1.02 to 1.87) | 0.036 |
| 9 | 1.26 | Non-CVD/cancer mortality | 1.15 (0.84 to 1.58) | 0.377 |
| 10 | 1.41 | Non-CVD/cancer mortality | 0.90 (0.67 to 1.22) | 0.506 |

a. ApoB, apolipoprotein B; CAD, coronary artery disease; CVD, cardiovascular disease.

b. Estimates are expressed as odds ratio (95% confidence interval) for CAD, or hazard ratio (95% confidence interval) for mortality outcomes per standard deviation increase in apoB.

**eTable 8. Stratum-specific Mendelian randomization estimates for genetically-predicted LDL-C on CAD, all-cause mortality, and cause-specific mortality.**

| Stratum | Mean LDL-C (mmol/L) | Outcome | Odds ratio/Hazard ratio | *P* value |
|---|---|---|---|---|
| 1 | 2.35 | CAD | 2.39 (2.12 to 2.70) | <0.001 |
| 2 | 2.74 | CAD | 2.35 (2.03 to 2.71) | <0.001 |
| 3 | 3.01 | CAD | 2.18 (1.83 to 2.58) | <0.001 |
| 4 | 3.24 | CAD | 2.31 (1.90 to 2.80) | <0.001 |
| 5 | 3.45 | CAD | 1.34 (1.09 to 1.65) | 0.005 |
| 6 | 3.65 | CAD | 1.81 (1.45 to 2.25) | <0.001 |
| 7 | 3.86 | CAD | 1.31 (1.04 to 1.66) | 0.023 |
| 8 | 4.10 | CAD | 1.52 (1.21 to 1.91) | <0.001 |
| 9 | 4.41 | CAD | 1.23 (0.98 to 1.56) | 0.077 |
| 10 | 4.90 | CAD | 1.34 (1.08 to 1.67) | 0.008 |
| 1 | 2.35 | All-cause mortality | 1.10 (0.96 to 1.25) | 0.171 |
| 2 | 2.74 | All-cause mortality | 1.22 (1.05 to 1.42) | 0.011 |
| 3 | 3.01 | All-cause mortality | 1.24 (1.05 to 1.46) | 0.013 |
| 4 | 3.24 | All-cause mortality | 1.30 (1.09 to 1.55) | 0.003 |
| 5 | 3.45 | All-cause mortality | 1.07 (0.90 to 1.28) | 0.433 |
| 6 | 3.65 | All-cause mortality | 1.30 (1.09 to 1.55) | 0.004 |
| 7 | 3.86 | All-cause mortality | 1.09 (0.91 to 1.31) | 0.347 |
| 8 | 4.10 | All-cause mortality | 1.04 (0.87 to 1.25) | 0.636 |
| 9 | 4.41 | All-cause mortality | 1.14 (0.95 to 1.37) | 0.169 |
| 10 | 4.90 | All-cause mortality | 0.99 (0.83 to 1.17) | 0.885 |
| 1 | 2.35 | CVD mortality | 1.64 (1.26 to 2.15) | <0.001 |
| 2 | 2.74 | CVD mortality | 1.87 (1.36 to 2.57) | <0.001 |
| 3 | 3.01 | CVD mortality | 1.43 (1.00 to 2.07) | 0.053 |
| 4 | 3.24 | CVD mortality | 2.00 (1.36 to 2.94) | <0.001 |
| 5 | 3.45 | CVD mortality | 0.87 (0.58 to 1.30) | 0.496 |
| 6 | 3.65 | CVD mortality | 2.10 (1.39 to 3.16) | <0.001 |
| 7 | 3.86 | CVD mortality | 1.21 (0.80 to 1.82) | 0.372 |
| 8 | 4.10 | CVD mortality | 1.35 (0.88 to 2.08) | 0.174 |
| 9 | 4.41 | CVD mortality | 1.57 (1.00 to 2.47) | 0.051 |
| 10 | 4.90 | CVD mortality | 1.15 (0.78 to 1.71) | 0.478 |
| 1 | 2.35 | Cancer mortality | 0.87 (0.71 to 1.06) | 0.167 |
| 2 | 2.74 | Cancer mortality | 1.07 (0.86 to 1.33) | 0.556 |
| 3 | 3.01 | Cancer mortality | 1.17 (0.92 to 1.48) | 0.194 |
| 4 | 3.24 | Cancer mortality | 1.17 (0.92 to 1.49) | 0.210 |
| 5 | 3.45 | Cancer mortality | 1.23 (0.97 to 1.57) | 0.094 |
| 6 | 3.65 | Cancer mortality | 1.04 (0.82 to 1.33) | 0.730 |
| 7 | 3.86 | Cancer mortality | 1.01 (0.79 to 1.30) | 0.920 |
| 8 | 4.10 | Cancer mortality | 0.96 (0.76 to 1.23) | 0.770 |
| 9 | 4.41 | Cancer mortality | 0.98 (0.77 to 1.26) | 0.891 |
| 10 | 4.90 | Cancer mortality | 0.96 (0.76 to 1.20) | 0.712 |

| | | | | |
|---|---|---|---|---|
| 1 | 2.35 | Non-CVD/cancer mortality | 1.10 (0.87 to 1.40) | 0.425 |
| 2 | 2.74 | Non-CVD/cancer mortality | 1.05 (0.79 to 1.40) | 0.719 |
| 3 | 3.01 | Non-CVD/cancer mortality | 1.14 (0.84 to 1.56) | 0.406 |
| 4 | 3.24 | Non-CVD/cancer mortality | 1.17 (0.84 to 1.61) | 0.349 |
| 5 | 3.45 | Non-CVD/cancer mortality | 0.94 (0.67 to 1.31) | 0.702 |
| 6 | 3.65 | Non-CVD/cancer mortality | 1.49 (1.04 to 2.13) | 0.031 |
| 7 | 3.86 | Non-CVD/cancer mortality | 1.21 (0.84 to 1.75) | 0.302 |
| 8 | 4.10 | Non-CVD/cancer mortality | 1.02 (0.72 to 1.46) | 0.892 |
| 9 | 4.41 | Non-CVD/cancer mortality | 1.29 (0.88 to 1.88) | 0.188 |
| 10 | 4.90 | Non-CVD/cancer mortality | 0.99 (0.69 to 1.41) | 0.945 |

a. CAD, coronary artery disease; CVD, cardiovascular disease; LDL-C, low-density lipoprotein cholesterol.

b. Estimates are expressed as odds ratio (95% confidence interval) for CAD, or hazard ratio (95% confidence interval) for mortality outcomes per standard deviation increase in LDL-C.

**eTable 9. Stratum-specific Mendelian randomization estimates for genetically-predicted TG on CAD, all-cause mortality, and cause-specific mortality.**

| Stratum | Mean TG (mmol/L) | Outcome | Odds ratio/Hazard ratio | *P* value |
|---|---|---|---|---|
| 1 | 0.76 | CAD | 8.76 (6.15 to 12.47) | <0.001 |
| 2 | 0.95 | CAD | 3.46 (2.53 to 4.74) | <0.001 |
| 3 | 1.11 | CAD | 2.93 (2.18 to 3.95) | <0.001 |
| 4 | 1.27 | CAD | 2.79 (2.10 to 3.70) | <0.001 |
| 5 | 1.44 | CAD | 1.82 (1.38 to 2.40) | <0.001 |
| 6 | 1.64 | CAD | 1.57 (1.21 to 2.05) | 0.001 |
| 7 | 1.87 | CAD | 1.58 (1.22 to 2.05) | 0.001 |
| 8 | 2.17 | CAD | 1.38 (1.08 to 1.76) | 0.011 |
| 9 | 2.62 | CAD | 1.34 (1.04 to 1.73) | 0.022 |
| 10 | 3.51 | CAD | 1.15 (0.91 to 1.45) | 0.237 |
| 1 | 0.76 | All-cause mortality | 2.78 (2.06 to 3.76) | <0.001 |
| 2 | 0.95 | All-cause mortality | 0.86 (0.65 to 1.14) | 0.291 |
| 3 | 1.11 | All-cause mortality | 1.26 (0.96 to 1.65) | 0.092 |
| 4 | 1.27 | All-cause mortality | 1.19 (0.92 to 1.55) | 0.187 |
| 5 | 1.44 | All-cause mortality | 1.10 (0.85 to 1.42) | 0.474 |
| 6 | 1.64 | All-cause mortality | 1.15 (0.89 to 1.47) | 0.283 |
| 7 | 1.87 | All-cause mortality | 1.08 (0.84 to 1.39) | 0.536 |
| 8 | 2.17 | All-cause mortality | 1.04 (0.82 to 1.31) | 0.770 |
| 9 | 2.62 | All-cause mortality | 1.05 (0.82 to 1.34) | 0.696 |
| 10 | 3.51 | All-cause mortality | 0.99 (0.79 to 1.24) | 0.905 |
| 1 | 0.76 | CVD mortality | 2.37 (1.18 to 4.75) | 0.015 |
| 2 | 0.95 | CVD mortality | 0.75 (0.40 to 1.42) | 0.380 |
| 3 | 1.11 | CVD mortality | 3.27 (1.76 to 6.07) | <0.001 |
| 4 | 1.27 | CVD mortality | 1.13 (0.64 to 2.01) | 0.675 |
| 5 | 1.44 | CVD mortality | 1.40 (0.79 to 2.47) | 0.253 |
| 6 | 1.64 | CVD mortality | 1.37 (0.79 to 2.38) | 0.255 |
| 7 | 1.87 | CVD mortality | 1.37 (0.79 to 2.35) | 0.259 |
| 8 | 2.17 | CVD mortality | 1.03 (0.62 to 1.71) | 0.896 |
| 9 | 2.62 | CVD mortality | 1.17 (0.68 to 1.99) | 0.573 |
| 10 | 3.51 | CVD mortality | 0.99 (0.62 to 1.60) | 0.981 |
| 1 | 0.76 | Cancer mortality | 2.57 (1.67 to 3.96) | <0.001 |
| 2 | 0.95 | Cancer mortality | 0.56 (0.39 to 0.83) | 0.003 |
| 3 | 1.11 | Cancer mortality | 1.01 (0.70 to 1.47) | 0.949 |
| 4 | 1.27 | Cancer mortality | 1.06 (0.73 to 1.52) | 0.768 |
| 5 | 1.44 | Cancer mortality | 1.01 (0.71 to 1.44) | 0.951 |
| 6 | 1.64 | Cancer mortality | 1.10 (0.78 to 1.56) | 0.591 |
| 7 | 1.87 | Cancer mortality | 0.99 (0.70 to 1.42) | 0.977 |
| 8 | 2.17 | Cancer mortality | 1.02 (0.73 to 1.42) | 0.912 |
| 9 | 2.62 | Cancer mortality | 0.97 (0.69 to 1.35) | 0.852 |
| 10 | 3.51 | Cancer mortality | 1.04 (0.75 to 1.44) | 0.819 |

| | | | | |
|---|---|---|---|---|
| 1 | 0.76 | Non-CVD/cancer mortality | 3.50 (2.05 to 5.97) | <0.001 |
| 2 | 0.95 | Non-CVD/cancer mortality | 2.18 (1.29 to 3.69) | 0.004 |
| 3 | 1.11 | Non-CVD/cancer mortality | 1.08 (0.65 to 1.81) | 0.767 |
| 4 | 1.27 | Non-CVD/cancer mortality | 1.55 (0.95 to 2.54) | 0.079 |
| 5 | 1.44 | Non-CVD/cancer mortality | 1.14 (0.69 to 1.88) | 0.603 |
| 6 | 1.64 | Non-CVD/cancer mortality | 1.08 (0.67 to 1.73) | 0.760 |
| 7 | 1.87 | Non-CVD/cancer mortality | 1.11 (0.68 to 1.83) | 0.676 |
| 8 | 2.17 | Non-CVD/cancer mortality | 1.06 (0.67 to 1.69) | 0.801 |
| 9 | 2.62 | Non-CVD/cancer mortality | 1.15 (0.70 to 1.86) | 0.582 |
| 10 | 3.51 | Non-CVD/cancer mortality | 0.88 (0.57 to 1.37) | 0.579 |

a. CAD, coronary artery disease; CVD, cardiovascular disease; TG, triglycerides.

b. Estimates are expressed as odds ratio (95% confidence interval) for CAD, or hazard ratio (95% confidence interval) for mortality outcomes per standard deviation increase in TG.

**eTable 10. Non-linear Mendelian randomization tests for genetically-predicted apoB, LDL-C, and TG on CAD, all-cause mortality, and cause-specific mortality overall, by sex, and by age.**

| Exposure | Outcome | Subgroup | *P* value (trend test) | *P* value (fractional polynomial test) |
|---|---|---|---|---|
| ApoB | CAD | Overall | <0.001 | <0.001 |
| | CAD | Men | <0.001 | <0.001 |
| | CAD | Women | 0.076 | 0.078 |
| | CAD | <65 years | <0.001 | <0.001 |
| | CAD | >=65 years | <0.001 | <0.001 |
| | All-cause mortality | Overall | 0.139 | 0.167 |
| | All-cause mortality | Men | 0.572 | 0.598 |
| | All-cause mortality | Women | 0.147 | 0.235 |
| | All-cause mortality | <65 years | 0.905 | >0.999 |
| | All-cause mortality | >=65 years | 0.088 | 0.236 |
| | CVD mortality | Overall | 0.438 | 0.437 |
| | CVD mortality | Men | 0.435 | 0.282 |
| | CVD mortality | Women | 0.314 | 0.442 |
| | CVD mortality | <65 years | 0.402 | 0.427 |
| | CVD mortality | >=65 years | 0.279 | 0.238 |
| | Cancer mortality | Overall | 0.366 | 0.590 |
| | Cancer mortality | Men | 0.809 | 0.916 |
| | Cancer mortality | Women | 0.418 | 0.634 |
| | Cancer mortality | <65 years | 0.963 | 0.827 |
| | Cancer mortality | >=65 years | 0.494 | 0.898 |
| | Non-CVD/cancer mortality | Overall | 0.875 | 0.926 |
| | Non-CVD/cancer mortality | Men | 0.635 | 0.638 |
| | Non-CVD/cancer mortality | Women | 0.556 | 0.766 |
| | Non-CVD/cancer mortality | <65 years | 0.191 | 0.551 |
| | Non-CVD/cancer mortality | >=65 years | 0.458 | 0.603 |
| LDL-C | CAD | Overall | <0.001 | <0.001 |
| | CAD | Men | <0.001 | <0.001 |
| | CAD | Women | 0.001 | 0.012 |
| | CAD | <65 years | <0.001 | <0.001 |
| | CAD | >=65 years | <0.001 | <0.001 |
| | All-cause mortality | Overall | 0.035 | 0.110 |
| | All-cause mortality | Men | 0.749 | >0.999 |
| | All-cause mortality | Women | 0.031 | 0.143 |
| | All-cause mortality | <65 years | 0.161 | 0.356 |
| | All-cause mortality | >=65 years | 0.017 | 0.054 |
| | CVD mortality | Overall | 0.095 | 0.114 |
| | CVD mortality | Men | 0.309 | 0.277 |
| | CVD mortality | Women | 0.343 | 0.476 |
| | CVD mortality | <65 years | 0.043 | 0.130 |

| | | | | |
|---|---|---|---|---|
| | CVD mortality | >=65 years | 0.182 | 0.212 |
| | Cancer mortality | Overall | 0.294 | 0.645 |
| | Cancer mortality | Men | 0.945 | >0.999 |
| | Cancer mortality | Women | 0.248 | 0.449 |
| | Cancer mortality | <65 years | 0.539 | 0.813 |
| | Cancer mortality | >=65 years | 0.277 | 0.540 |
| | Non-CVD/cancer mortality | Overall | 0.823 | >0.999 |
| | Non-CVD/cancer mortality | Men | 0.360 | 0.380 |
| | Non-CVD/cancer mortality | Women | 0.164 | 0.542 |
| | Non-CVD/cancer mortality | <65 years | 0.657 | 0.774 |
| | Non-CVD/cancer mortality | >=65 years | 0.191 | 0.281 |
| TG | CAD | Overall | <0.001 | <0.001 |
| | CAD | Men | <0.001 | <0.001 |
| | CAD | Women | <0.001 | <0.001 |
| | CAD | <65 years | <0.001 | <0.001 |
| | CAD | >=65 years | <0.001 | <0.001 |
| | All-cause mortality | Overall | 0.007 | 0.003 |
| | All-cause mortality | Men | 0.257 | 0.079 |
| | All-cause mortality | Women | 0.248 | 0.075 |
| | All-cause mortality | <65 years | 0.177 | 0.244 |
| | All-cause mortality | >=65 years | 0.215 | 0.121 |
| | CVD mortality | Overall | 0.006 | 0.010 |
| | CVD mortality | Men | 0.026 | 0.036 |
| | CVD mortality | Women | 0.165 | 0.148 |
| | CVD mortality | <65 years | 0.440 | 0.376 |
| | CVD mortality | >=65 years | 0.003 | 0.005 |
| | Cancer mortality | Overall | 0.813 | 0.755 |
| | Cancer mortality | Men | 0.282 | 0.441 |
| | Cancer mortality | Women | 0.607 | 0.641 |
| | Cancer mortality | <65 years | 0.849 | >0.999 |
| | Cancer mortality | >=65 years | 0.042 | 0.046 |
| | Non-CVD/cancer mortality | Overall | 0.001 | 0.002 |
| | Non-CVD/cancer mortality | Men | 0.081 | 0.069 |
| | Non-CVD/cancer mortality | Women | 0.021 | 0.012 |
| | Non-CVD/cancer mortality | <65 years | 0.003 | 0.074 |
| | Non-CVD/cancer mortality | >=65 years | 0.025 | 0.027 |

a. ApoB, apolipoprotein B; CAD, coronary artery disease; CVD, cardiovascular disease; LDL-C, low-density lipoprotein cholesterol; TG, triglycerides.

b. The trend test assesses whether a linear trend in the stratum-specific estimates exists, and the fractional polynomial test examines whether a non-linear model fits the exposure-outcome relationship better than a linear model.
